# A complex duplication overlapping *FBRSL1* implicated in a developmental and epileptic encephalopathy

**DOI:** 10.64898/2026.03.30.26349353

**Authors:** Lital Cohen-Vig, Jacob E Munro, Joshua Reid, Tom Witkowski, Neblina Sikta, Dror Kraus, Mark F Bennett, Ingrid E Scheffer, Michael S Hildebrand, Melanie Bahlo, Samuel F Berkovic

**Affiliations:** Department of Medicine, Epilepsy Research Centre, University of Melbourne, Austin Health, Heidelberg, Victoria, Australia; Department of Pediatric Neurology, Schneider Children’s Medical Center of Israel, Petah Tikva, Israel; Genetics and Gene Regulation Division, The Walter and Eliza Hall Institute of Medical Research, Parkville, Victoria, Australia; Department of Medical Biology, The University of Melbourne, Parkville, Victoria, Australia; Gray Faculty of Medicine and Health Sciences, Tel-Aviv University, Tel Aviv 6997801, Israel; Florey Institute of Neuroscience and Mental Health, Heidelberg, Victoria, Australia; Department of Pediatrics, University of Melbourne, Melbourne, Victoria, Australia; Murdoch Children’s Research Institute, Parkville, Victoria, Australia

**Keywords:** Contractures, Developmental and epileptic encephalopathy, Feeding difficulties, *FBRSL1* gene duplication, Respiratory insufficiency, Structural variants

## Abstract

To date, *FBRSL1*-related disorder has been reported in five individuals with congenital abnormalities and severe postnatal impairment with or without epilepsy; however, the full extent of the phenotypic and genotypic spectrum remains unclear. Previously reported cases involved small truncating variants apparently escaping nonsense-mediated decay, suggesting either a haploinsufficiency or a dominant-negative mechanism. We report the first case of a complex structural variant at the *FBRSL1* locus, resulting in an additional, partially truncated copy of the gene, providing strong evidence for a dominant-negative mechanism. RNA-Seq supported the expression of the additional truncated gene copy.

The patient is an infant girl with a profound developmental and epileptic encephalopathy (DEE). The child presented at birth with intrauterine growth restriction, respiratory insufficiency, severe swallowing dysfunction, spasticity, contractures, optic nerve hypoplasia, facial dysmorphism, and atrial septal defect. She developed severe postnatal growth restriction with microcephaly and profound developmental impairment. She has a DEE with frequent neonatal focal seizures evolving to infantile epileptic spasms syndrome (IESS). Our patient has congenital abnormalities in common with previously reported cases, along with a profound DEE, not associated previously with *FBRSL1*. Our case expands both the phenotypic and genotypic spectrum of *FBRSL1*-related disorder.

## Introduction

Developmental and epileptic encephalopathies (DEEs) are the most severe group of epilepsies, where developmental impairment is the result of both the underlying etiology and epileptic encephalopathy (Zuberi, Wirrell et al. 2022). The etiologies of DEEs are diverse, including genetic, structural, metabolic, infectious, immune, or idiopathic (Zuberi, Wirrell et al. 2022; Scheffer, Zuberi et al. 2024). Genetic testing of the epilepsies has the highest yield in the DEEs, with over 900 of the known 1000 monogenic epilepsy genes implicated in DEEs (Oliver, Scheffer et al. 2023). Pathogenic variants are currently detected in ∼ 50% of patients, of which 60-80% are single nucleotide variants, while 10-20% are deletions and duplications (Hebbar and Mefford 2020).

*FBRSL1* was first associated with human disease in 2020 in three children with congenital malformations (cardiac, cleft palate, gastrointestinal), global severe developmental delay, respiratory insufficiency, feeding difficulties, failure to thrive, acquired microcephaly, contractures, and facial dysmorphism (Ufartes, Berger et al. 2020). A fourth and fifth patient were subsequently reported with similar features as well as epilepsy (Bukvic, De Rinaldis et al. 2024; Xu, Zhang et al. 2026). All five patients had *FBRSL1* variants predicted to lead to protein truncation (Ufartes, Berger et al. 2020; Bukvic, De Rinaldis et al. 2024).

*FBRSL1* is a paralog of *AUTS2* which is associated with intellectual disability and microcephaly (Pauli, Berger et al. 2021). *FBRSL1*, and sister genes *AUTS2* and *FBRS,* encode proteins with shared conserved domains (Sellers, Robertson et al. 2020) and together form a tripartite gene family known as the AUTS2 family. *FBRSL1* is thought to be an RNA-binding protein (Baltz, Munschauer et al. 2012). *FBRSL1* and *AUTS2* encode proteins that form part of the Polycomb subcomplexes PRC 1.5 and PRC 1.3 and play an important role in embryonic development by regulating gene inactivation (Pauli, Berger et al. 2021). In animal models (zebrafish and *Xenopus laevis*), *FBRSL1* has been shown to be expressed and play a role in embryonic brain development, as well as in the development of the spinal cord, cranial nerves, somites, branchial arches, and the heart (Kondrychyn, Robra et al. 2017; Ufartes, Berger et al. 2020; Berger, Gerstner et al. 2024). Rescue of brain, craniofacial, and cardiac embryonic malformations has been achieved in *FBRSL1* animal models by overexpressing the short N-terminal *FBRSL1* transcript. These animal models support the role of *FBRSL1* in human embryonic development, with loss-of-function recapitulating a congenital malformation syndrome along with neurological features (Ufartes, Berger et al. 2020; Berger, Gerstner et al. 2024; Bukvic, De Rinaldis et al. 2024).

Herein we present the sixth patient with a germline disease-causing variant in *FBRSL1*, expanding the phenotypic spectrum to encompass DEE, and the genotypic spectrum to include partial gene duplication, inconsistent with haploinsufficiency. We emphasize the importance of a comprehensive rare variant analysis including SVs, as well as detailed phenotypic matching to prior case reports, in identifying new and emerging rare disease variant associations.

## Materials and Methods

### Patient Recruitment

The study was approved by the Austin Health Human Research Ethics Committee, Austin Health, Melbourne, Australia (study ID H2007/02961) and the WEHI Human Research Ethics Committee (project G20/01). The proband’s parents provided informed consent for participation in the study.

### GS Reanalysis

GS data (Illumina 151 bp paired-end sequencing) was processed following the GATK Best Practices guidelines for germline short variant discovery (Van der Auwera, Carneiro et al. 2013). Briefly, paired reads were aligned to the GRCh38 reference assembly using BWA-MEM (v0.7.17-r1188), following which GATK (v4.1.9) was used for base quality score recalibration (GATK BaseRecalibrator), germline short variant discovery (GATK HaplotypeCaller), joint variant calling (GATK GenotypeGVCFs) and variant quality score recalibration and filtering (GATK VariantRecalibrator and GATK ApplyVQSR). Germline variants were annotated with VEP (v122) (Hunt, Moore et al. 2022) and filtered for variants in known epilepsy genes (Genes4Epilepsy) (Oliver, Scheffer et al. 2023) and later for variants in all genes that are either de-novo or autosomal recessive. ExpansionHunter (v5.0.0) (Dolzhenko, Deshpande et al. 2019) and exSTRa (v1.1.0) (Tankard, Bennett et al. 2018) were used with default parameters to search for pathogenic expansions of known, disease-causing tandem repeats. Resulting candidate variants were then reviewed to exclude false-positive calls by inspection with IGV (Thorvaldsdóttir, Robinson et al. 2013).

SVs were called from the BAM files with the read-depth based caller CNVnator (v0.4.1) (Abyzov, Urban et al. 2011) along with the split-read/read-pair based callers Manta (v1.6.0) (Chen, Schulz-Trieglaff et al. 2016) and smoove (v0.2.5) (Pedersen BS 2020), with only variant calls supported by multiple SV callers pursued further. Germline short variants (SNPs/indels) along with SVs were annotated with VEP (v122) (Hunt, Moore et al. 2022) with the StructuralVariantOverlap plugin to annotate population allele frequencies from the gnomAD SV (v2.1) dataset. Rare variants matching recessive (max population allele frequency=0.01) and *de novo* (max population allele frequency=0.0001) inheritance patterns with a VEP impact of “MODERATE” or “HIGH” were extracted and intersected firstly with known epilepsy genes (Genes4Epilepsy) (Oliver, Scheffer et al. 2023) and subsequently for clinically relevant known mendelian disease genes (PanelApp Australia “Mendeliome”, v1.2063) (Martin, Williams et al. 2019). Resulting candidate variants were then manually reviewed to exclude false-positive calls with SVPV (Munro, Dunwoodie et al. 2017).

### SV Validation

#### GS analysis

Additional evidence to support and refine the complex duplication was derived from the short-read GS data. Sequencing coverage for the trio GS on the entirety of the affected chromosome was extracted with mosdepth (Pedersen and Quinlan 2018) in 500 bp bins, and then normalized by dividing by the median non-zero coverage. The proband’s coverage was then further normalized by dividing by the mean normalized coverage of the unaffected parents in each bin to generate an estimate of copy number across the complex SV region. Additionally, the variant allele frequencies of SNVs were extracted from the GATK short-variant callset and assigned as either maternal, paternal or ambiguous origin allowing determination of which allele (maternal or paternal) carried the *de novo* SV.

#### Droplet Digital PCR (ddPCR)

DNA was extracted from 300 µl whole blood samples using Qiagen Puregene blood kit as per the manufacturer’s instructions (Puregene Handbook 01/2022). The 100 µl DNA elution was then sheared using a Covaris g-TUBE as per the manufacturer’s guidelines. Custom primers were ordered from Bioneer Pacific (Kew East, Australia) to validate and quantify the SV (primer sequences are available on request). Droplet generation, PCR cycling, and droplet reading were performed according to the manufacturer’s recommendations (Bio-Rad, Hercules, CA). Primers were mixed with QX200™ ddPCR™ EvaGreen Supermix at a final concentration 0.1μM, and mixed with 10 ng of DNA sample to a final volume of 22 μL. Twenty microliters of reactions were loaded in an eight-channel droplet generator cartridge (Bio-Rad) and droplets were generated with 70 μL of droplet generation oil (Bio-Rad) using the manual QX200 Droplet Generator. Following droplet generation, samples were manually transferred to a 96-well PCR plate, heat-sealed and amplified on a C1000 Touch thermal cycler using the following cycling conditions: 95°C for 5 min for one cycle, followed by 40 cycles at 94°C for 30 s and 63°C for 60 s, one cycle at 4°C for 5 minutes, 90°C for 5 min and 4°C hold. Post-PCR products were read on the QX200 droplet reader (Bio-Rad) and analysed using the QuantaSoft software.

#### Long Read DNA Sequencing

DNA was sequenced with a PromethION 2 Integrated from Oxford Nanopore Technologies (ONT). DNA libraries were prepared starting with 3 µg of DNA using the Ligation kit SQK-LSK114 and sequenced with R10.4.1 flow cells (FLO-PRO114M) as per the manufacturers’ specifications. Briefly, reads were base called using dorado (v0.7.0) (Technologies 2024) and aligned to the hg38 human reference genome using minimap2 (v2.24-r1122) (Li 2018). Variant calling was run via the Epi2Me Human Variation pipeline (v1.8.3) (Labs 2024), using Sniffles2 (v2.0.7) (Smolka, Paulin et al. 2024) to identify SVs.

### RNA Sequencing

RNA was extracted from 9 mL whole-blood collected in PAXgene RNA tubes using the PAXgene blood RNA kit as per the manufacturer’s instructions (PreAnalytix PAXgene Blood RNA Kit Handbook). Libraries for one proband and one healthy control were prepared from blood-derived RNA using the Illumina Stranded Total RNA Prep with Ribo-Zero Plus kit and sequenced on the NovaSeq platform. Paired-end, 151 base pair reads were processed using the nf-core/rnaseq pipeline (v3.15.0)(Ewels, Peltzer et al. 2020), using STAR (v2.7.10a) (Dobin, Davis et al. 2013) to align reads to the GRCh38 reference genome (Ensembl release 112) and salmon (v1.10.1) (Patro, Duggal et al. 2017) to quantify exon expression. *FBRSL1* exon expression counts were normalized per million reads (TPM), and a ratio of proband to healthy control expression was calculated for each exon. The Mann-Whitney U test was used to assess the statistical significance of the difference in distribution of expression ratio between the duplicated and non-duplicated exons.

## Results

An infant girl presented with apnea and severe swallowing difficulties from birth. The pregnancy was complicated by maternal COVID-19 infection in the first and third trimesters; with intrauterine growth retardation noted at the end of the second trimester. She was delivered at 38 weeks’ gestation by spontaneous vaginal delivery, weighing 2.34 kg, with Apgar scores of eight and nine at one and five minutes respectively. From birth, she required nasogastric feeding due to inability to handle secretions or milk. She also required intermittent nasal ventilatory support for central apnea. Examination findings included small auricles with overfolded helices, bilateral Simian creases, narrow feet, optic hypoplasia, truncal hypotonia, spastic quadriparesis and contractures. Echocardiography showed a moderate-sized atrial septal defect. She developed postnatal growth restriction, with postnatal microcephaly. Brain magnetic resonance imaging (MRI) showed mildly dysplastic lateral ventricles and dilated extra axial cerebrospinal fluid spaces (Figure 1). Family history was unremarkable.

**Figure 1.**
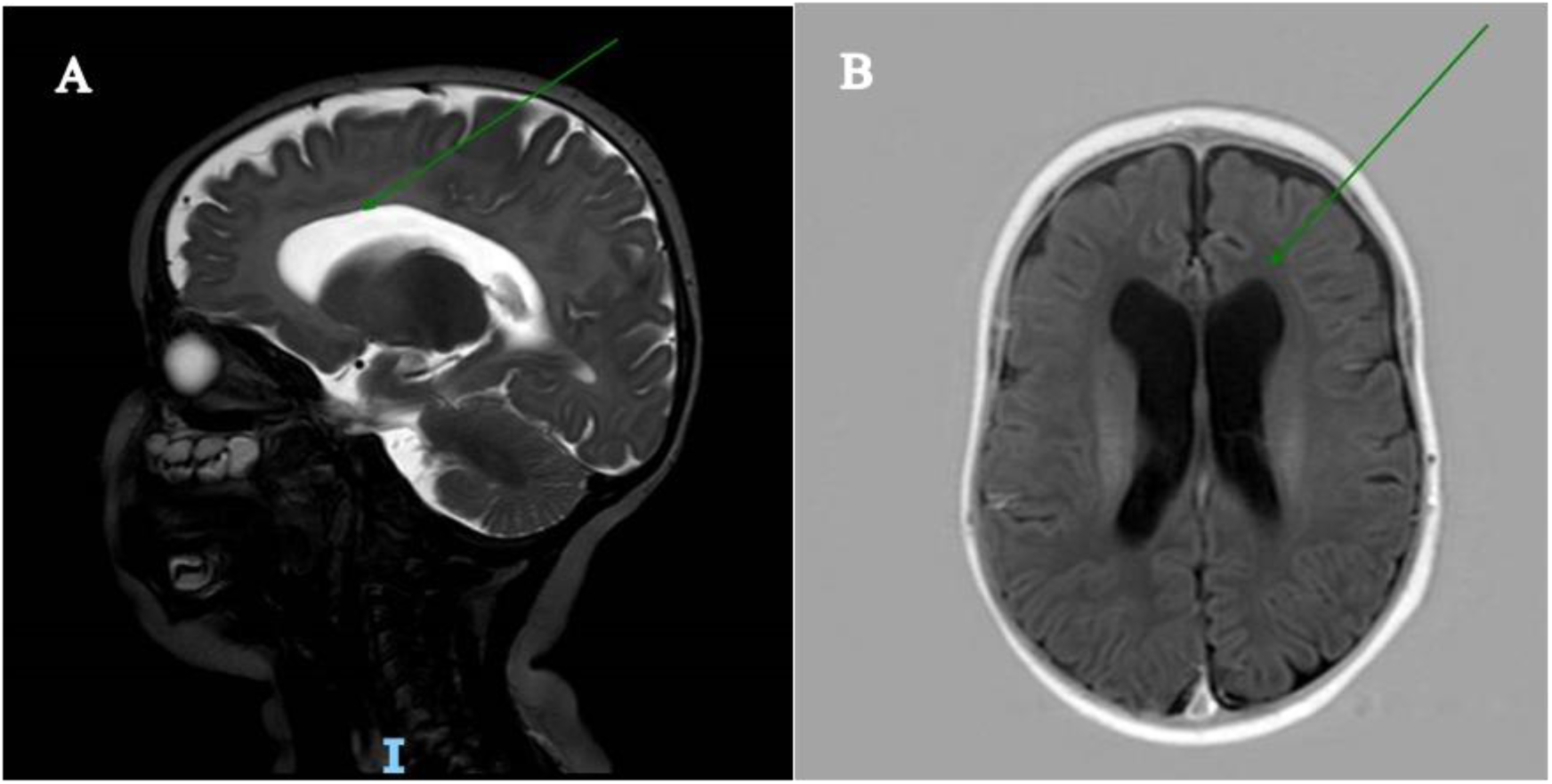
Magnetic Resonance Imaging (MRI); A-T2 weighted mid-sagittal brain imaging demonstrating excess extra-axial cerebrospinal fluid, and mildly dysplastic lateral ventricles (arrow). B-T1 weighted axial brain imaging demonstrating mildly dilated and dysplastic lateral ventricles (arrow).

From birth, she had frequent focal impaired consciousness seizures with prominent autonomic features including apnea and bradycardia. Between 3-6 months of age, focal clonic seizures developed. Both apneic seizures and clonic seizures markedly improved with high doses of phenobarbital (serum levels 45 mcg/mL, therapeutic range 15-40), and the ketogenic diet. After eight months, she developed epileptic spasms. Electroencephalogram (EEG) showed multifocal interictal epileptiform discharges that evolved to hypsarrhythmia before the age of 1 year. Vigabatrin was transiently effective, however, epileptic spasms and hypsarrhythmia recurred. High dose topiramate (25 mg/kg/day) was added (Glauser, Clark et al. 2000) to the treatment regimen of vigabatrin (185 mg/kg/day), phenobarbital (11 mg/kg/day), levetiracetam (60 mg/kg/day), and the ketogenic diet. Due to lack of efficacy, topiramate and levetiracetam were weaned. The addition of cannabis oil (up to 10 mg/kg/day) and clobazam (0.5 mg/kg/day) also proved ineffective. Valproate, lacosamide, and vitamin B6 were also ineffective. Severe global developmental delay was noted from birth, however temporary seizure control with the onset of hypsarrhythmia was associated with improved alertness which regressed at its recurrence 2 months later.

A genetic etiology was sought. Neonatal clinical trio GS was negative and research reanalysis of the data was performed through our research pipeline for all Mendelian disease-associated genes (PanelApp Australia Mendeliome (Martin, Williams et al. 2019)). We identified a *de novo* complex duplication on the paternal chromosome 12 haplotype (Figure 2). This duplication was predicted to have a breakpoint in the *FBRSL1* gene between exon 5 and 6 of the MANE Select transcript (NM_001367871.1, isoform 2 (Ufartes, Berger et al. 2020) (Figure 3). The phenotype was concordant with the five previous patients described with *FBRSL1* pathogenic variants (Table 1). Our patient has a neonatal onset, drug-resistant epilepsy associated with regression and profound developmental impairment, expanding the phenotypic spectrum of *FBRSL1* epilepsy to neonatal-onset DEE (Ufartes, Berger et al. 2020; Bukvic, De Rinaldis et al. 2024).

**Figure 2.**
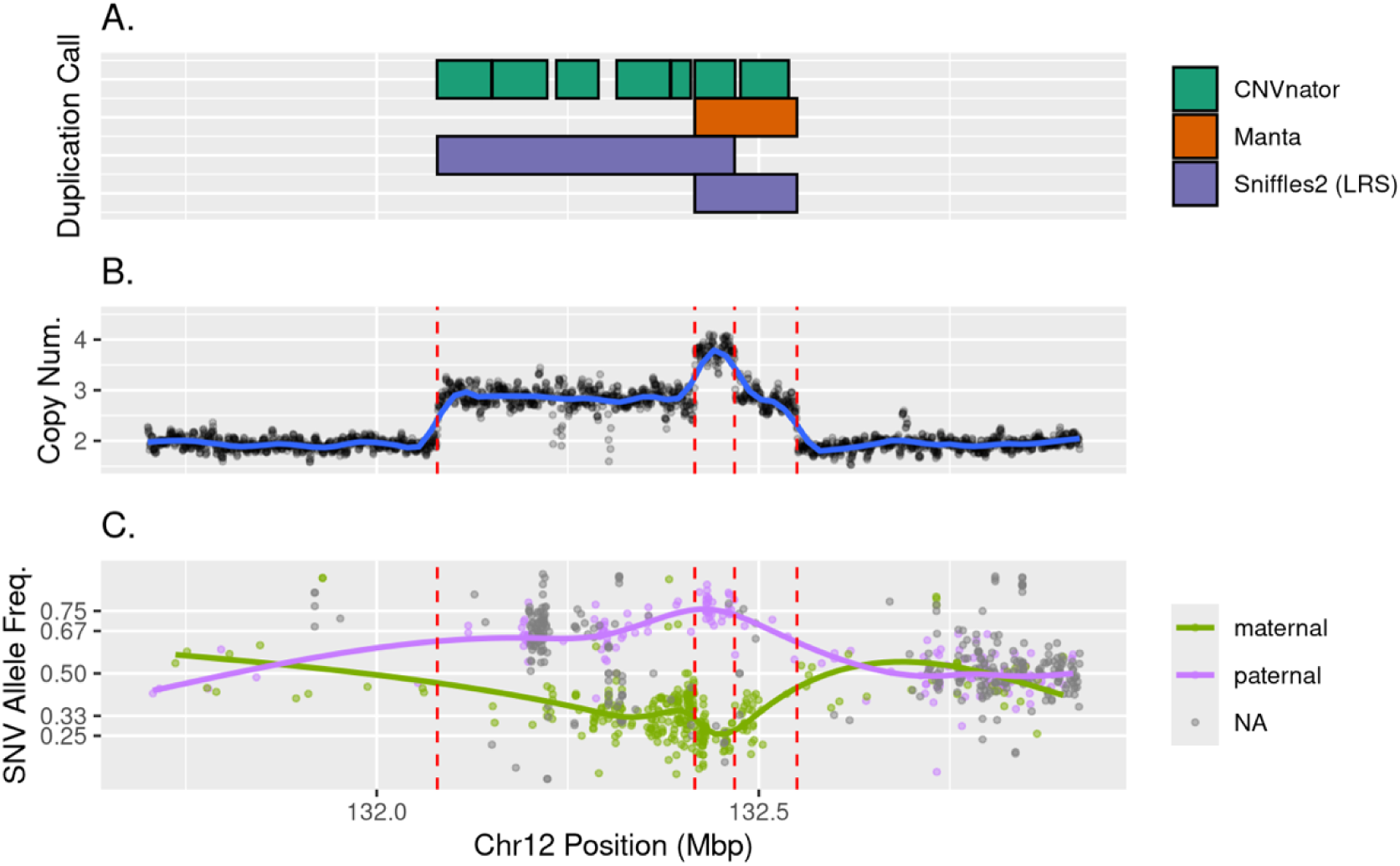
Evidence for complex FBRSL1 duplications from genome sequencing. A. Duplication variant calls on short-read data (CNVnator & Manta) and long-read data (Sniffles2). B. Normalised copy number in affected proband with local regression (LOESS) curve indicated by blue-line and duplication breakpoints are indicated by vertical dashed red lines. C. Variant-allele frequency of SNVs inherited maternally and paternally at the duplication region with local regression (LOESS) curves fitted.

**Figure 3.**
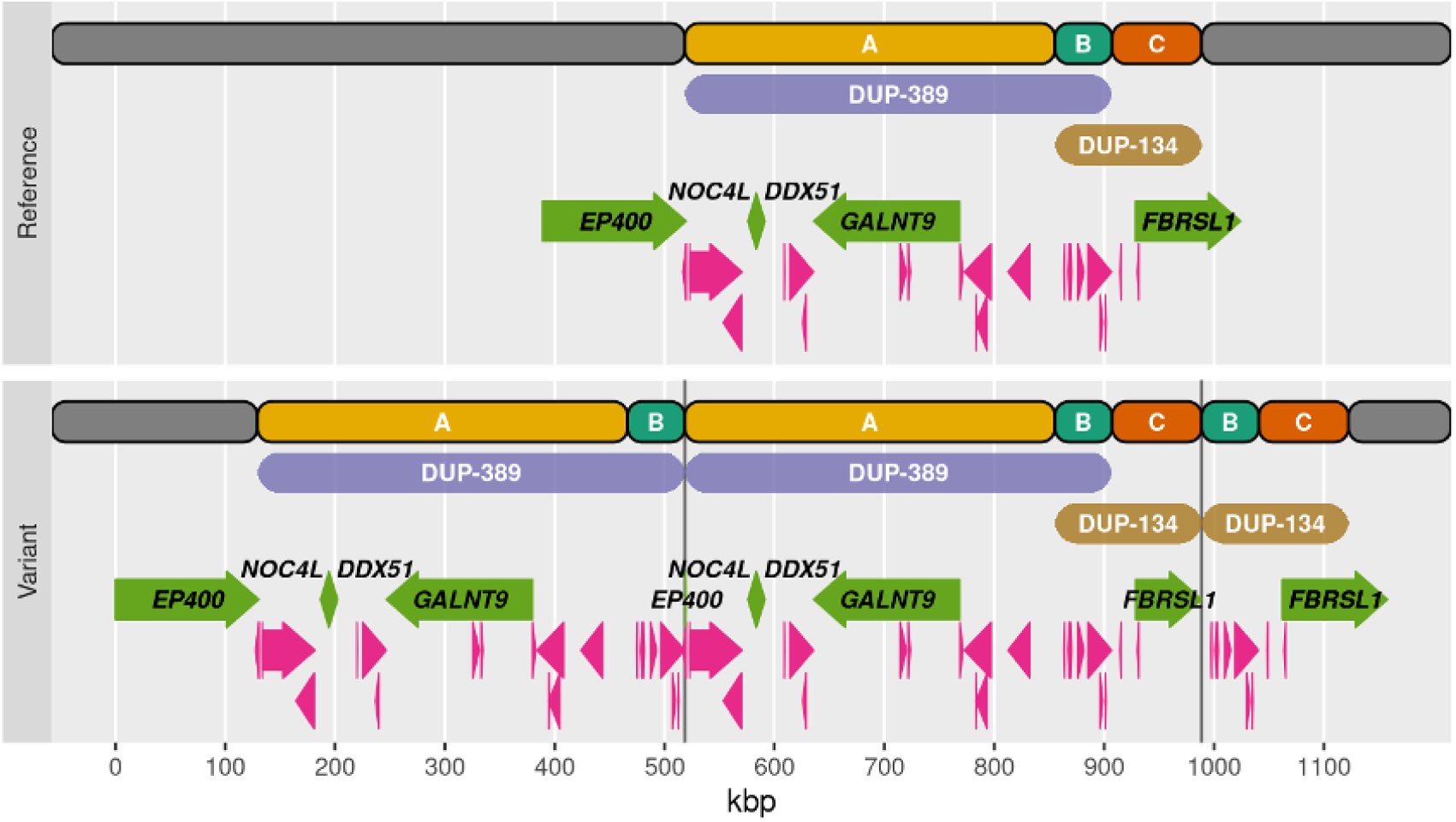
Schematic of the reference and complex structural variant alleles. The overlapping duplications DUP-389 and DUP-134 divide the reference region into three sections: A) with two copies in the variant allele; B) with three copies; and C) with two copies. Protein coding genes are represented as green arrows with labels, lncRNAs are represented as pink arrows (GENCODE v47 basic). Protein coding genes NOC4L, DDX51 and GALNT9 are fully duplicated. Vertical grey lines indicate the tandem duplication breakpoints giving rise to partial duplication of the protein coding genes FBRSL1 (promotor and majority of gene) and EP400 (3’UTR fragment only).

**Table 1:** Comparison of published phenotype of FBRSL1 patients (P1-5), and our patient’s (P6) phenotype. P1-3, described by Ufartes et al. (Ufartes, Berger et al. 2020); P4 described by Bukvic, et al.(Bukvic, De Rinaldis et al. 2024)); P5, described by Xu et al, (Xu, Zhang et al. 2026); P6 described by us herein; GDD, Global developmental delay; ID, Intellectual disability; ASD, Atrial septal defect; PDA, Patent ductus arteriosus; VSD, Ventricular septal defect;. † ID is so severe, it is not possible to define autistic behaviors separately; ‡ Skin creases were not observed in our patient when examined after 18 months, but in the other patients were most prominent in the first year of life and resolved with age; In bold some of the unique features, which are not prevalent in most DEEs; § profoundly delayed speech;

| Patient ID | P1 | P2 | P3 | P4 | P5 | P6 |
| --- | --- | --- | --- | --- | --- | --- |
| ***FBRSL1* variant type** | Short isoform truncating variant (confirmed NMD escape) | Short isoform truncating variant (confirmed NMD escape) | Short isoform truncating variant (confirmed NMD escape) | Short isoform truncating variant (untested NMD escape) | Short isoform truncating variant (untested NMD escape) | 5' partial duplication, spanning all short isoforms |
| **GDD/ ID** | + | + | + | + | + | + |
| **No speech** | + | + | + | + | § | + |
| **Autistic behavior** | + | + | + | + | + | Inconclusive † |
| **Microcephaly** | + | + | + | + | + | + |
| **Feeding difficulty** | + | + | + | + | - | + |
| **Failure to thrive** | + | + | + | + | + | + |
| **Contractures** | + | + | + | + | + | + |
| **Heart defects** | ASD/PDA | ASD/VSD | - | - | ASD | ASD |
| **Facial dysmorphism** | + | + | + | + | + | + |
| **Cleft palate** | + | + | - | - | - | - |
| **Respiratory insufficiency** | + | + | + | - | - | + |
| **Hearing impairment** | + | + | - | - | - | Inconclusive |
| **Skin creases** | + | + | - | - | - | Inconclusive ‡ |
| **Other malformations** | Anus abnormality | Asplenia | - | Hip dislocation | - | Optic nerve hypoplasia |
| **Seizures** | - | - | - | + | + | + |

It took several steps to validate the precise structure of this complex SV. Short-read SV callers did not clearly resolve the SV. Manta and Smoove predicted a single duplication of 134 kb (chr12:132416214-132549850, GRCh38) intersecting with *FBRSL1*, while CNVnator predicted seven separate duplications spanning a region of 460 kb (chr12:132079000-132539000, GRCh38) (Figure 2A). Further detailed analysis of the short-read data revealed that the variant was composed of two overlapping duplications (Figure 2B-C). We then used ddPCR to validate the duplication of the 5’ end of one allele of *FBRSL1*. Finally, ONT long-read sequencing analysis with the SV caller Sniffles2 (Smolka, Paulin et al. 2024) resolved the breakpoints of the two overlapping tandem duplications, one of 389 kb (DUP-389, chr12:132079305-132468226, GRCh38) and the second of 134 kb (DUP-134, chr12:132416214-132549850, GRCh38), with an 82 kb region included in both duplications (Figure 2A).

DUP-134, which was the only one to directly affect *FBRSL1*, covered a region beginning 74 kb upstream of the *FBRSL1* transcriptional start site and extended downstream over the first five exons of isoforms 1 (NM_001142641.2) and 2 (NM_001367871.1), and across all exons of isoforms 3.1 (NM_001382741.1) and 3.2 (ENST00000542061.2). The upstream region contained three additional transcripts, all of which were long non-coding RNAs of unknown function (ENSG00000256783, ENSG00000256875 and ENSG00000274373, GENCODE v43). As the promoter and transcriptional start site were both upstream of the *FBRSL1* breakpoint, it is likely that both the normal *FBRSL1* and the partially duplicated copy will be expressed. Expression of the partially duplicated copy is expected to extend into the intergenic region that lies upstream of the complete *FBRSL1* copy (Figure 3). Indeed, RNA sequencing on our patient’s blood confirmed increased expression of the duplicated *FBRSL1* exons (p<0.001, Mann–Whitney U test) and increased expression of the intergenic region upstream of *FBRSL1,* compared to a healthy control (Figure 4).

**Figure 4.**
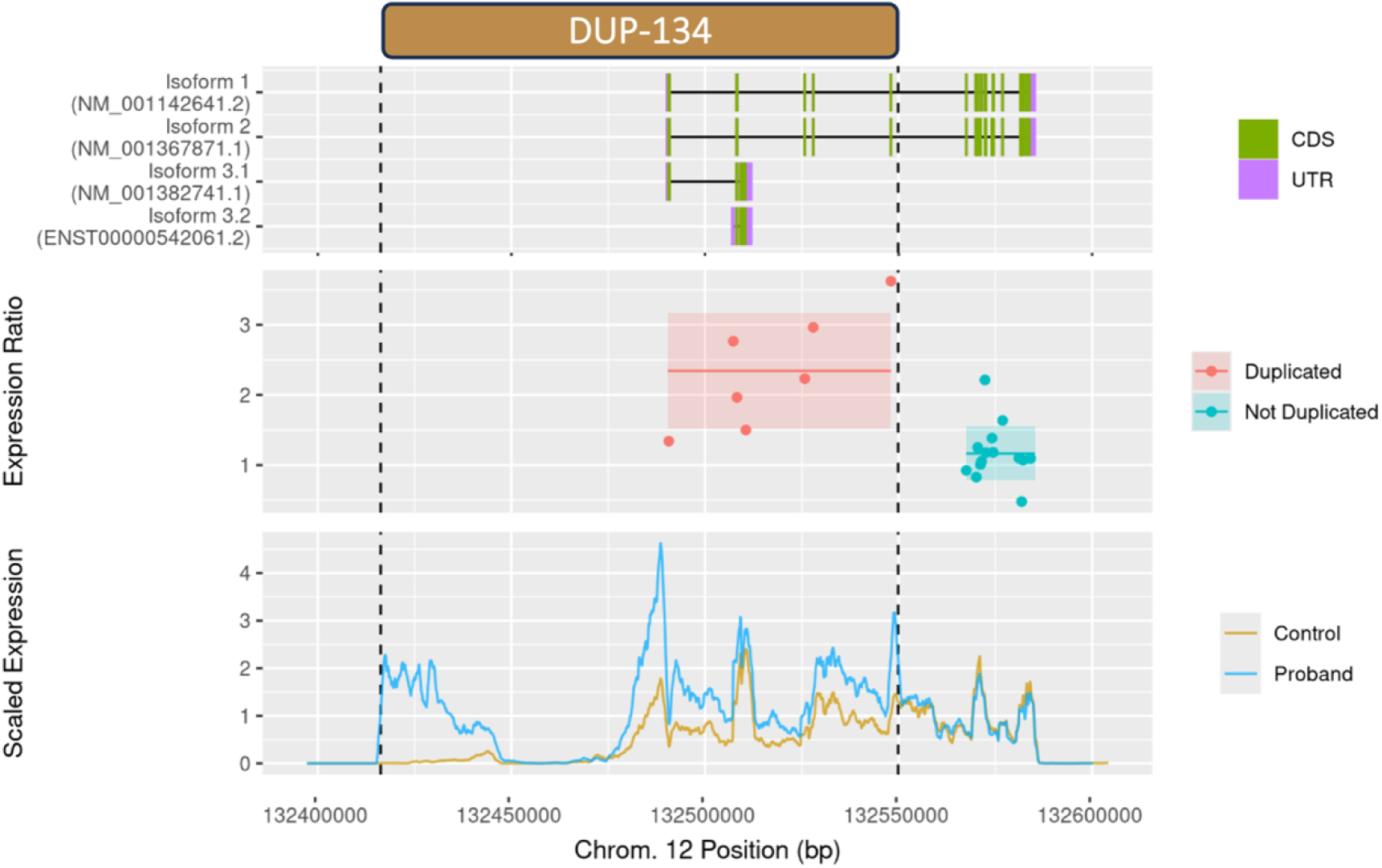
RNA expression of the FBRSL1 microduplication region. Dashed vertical lines indicate the breakpoints of the overlapping duplication (DUP-134). Top panel) Schematic showing coordinates of wild type FBRSL1 isoforms (oriented 5’–3’ on the sense strand). Middle panel) Exonic expression ratio of proband to control, showing higher expression of FBRSL1 exons in the duplicated region. The horizontal line indicates the mean in the duplicated region (red) and in the non-duplicated (teal), and the shaded rectangle indicates the range from +1 to -1 standard deviation. Bottom panel) RNA expression in the proband compared to healthy control across the entire FBRSL1 region. Expression levels are scaled by dividing by the mean expression in non-duplicated 3’ end of FBRSL1 within each sample. Expression is calculated in 100 bp bins and plotted as a moving average over 20 bins. Increased expression is observed in the proband in both the upstream region as well as the 5’ duplicated end of FBRSL1.bp base pair. CDS – coding DNA sequence; UTR – untranslated region;

## Discussion

We report an infant with profound impairment associated with a developmental and epileptic encephalopathy. She had a *de novo* complex SV comprising two duplications (DUP-389 and DUP-134) with an overlapping region. DUP-134 resulted in a partial duplication of *FBRSL1*, while DUP-389 resulted in the amplification of three protein coding genes (*DDX51*, *NOC4L*, *GALNT9*), as well as a portion of the 3’ untranslated region (UTR) of EP400. Several long noncoding RNAs (lncRNAs) lie within the complex SV region but none have previously been associated with disease.

Of the other affected protein coding genes, only *EP400* has been associated with neurodevelopmental delay and epilepsy in 11 patients (Luo, Wang et al. 2025). All had drug-responsive epilepsy, milder intellectual disability (8 mild, 1 moderate, 2 severe), with compound heterozygote *EP400* pathogenic variants. However, in our patient, the larger of the duplications (DUP-389) only involved the 3’ untranslated region (UTR) of *EP400*, and the phenotypic match with our patient was weak. Our patient had a range of features not seen in individuals with *EP400* pathogenic variants, which have been described in patients with *FBRSL1* related disorder, including respiratory insufficiency, swallowing difficulties, contractures, and heart malformations. In addition, *FBRSL1* variants are associated with neurodevelopmental disorders, with or without epilepsy (Ufartes, Berger et al. 2020; Bukvic, De Rinaldis et al. 2024; Xu, Zhang et al. 2026). Interestingly, its paralog *AUTS2* has also been associated with epilepsy (Biel, Castanza et al. 2022). Furthermore, the complex duplication is predicted to have a larger impact on *FBRSL1* than other genes due to the intronic breakpoint.

Our patient expands the phenotypic spectrum of *FBRSL1*-related disorders to include DEE. Previously described patients (P1-5) had congenital malformations and intellectual disability, similar to our patient. Two patients (P4, P5) had epilepsy. Their epilepsy was milder, with childhood onset approximately at the age of 4 years. P4’s epilepsy was initially controlled by valproate but then seizures reoccurred and only improved with additional ethosuximide and rufinamide treatment. P5 had drug responsive epilepsy. Neither had developmental regression (Ufartes, Berger et al. 2020; Bukvic, De Rinaldis et al. 2024; Xu, Zhang et al. 2026). Our patient (P6) has a severe neonatal-onset drug-resistant DEE. Our findings confirm the association of *FBRSL1* with epilepsy and expand its phenotypic spectrum to include severe, neonatal-onset, DEE. All five reported patients have truncating variants: two have nonsense variants, and three have frameshift variants leading to a premature stop codon. All truncating variants described affect the short N-terminal *FBRSL1* isoforms (3.1 (XM_005266181.4) and 3.2 (ENST00000542061.2)), which include a DNA translocase domain not present on the long isoforms 1 (NM_001142641.2) and 2 (NM_001367871.1)). Functional studies have shown that these truncations disrupt protein function and phenotypic rescue only occurs with the intact short N-terminal isoform. The molecular mechanism may be either haploinsufficiency of the short N-terminal isoforms or a dominant negative effect following escape from nonsense mediated decay (NMD) (Ufartes, Berger et al. 2020; Bukvic, De Rinaldis et al. 2024). NMD escape was experimentally confirmed for pathogenic variants of patients P1-3 (Ufartes, Berger et al. 2020).

The partial duplication of *FBRSL1* in our patient is not consistent with haploinsufficiency, since the patient retains two intact *FBRSL1* alleles. The duplication encompasses the entire short N terminal isoforms (3.1 and 3.2) and first 5 exons of the longer isoforms (1 and 2). There is no evidence that our patient’s complex duplication caused truncation of the short N-terminal isoforms. We observed increased expression of exons preceding the breakpoint, and read-through expression of the intergenic region upstream of *FBRSL1.* We postulate that pathogenicity arises from expression of the incomplete copy of *FBRSL1*, along with the read-through intergenic sequence, which is translated into an aberrant protein product that has a dominant negative effect on the normal short isoform potentially by competing for binding of molecular targets. There is limited understanding of the precise molecular function of *FBRSL1* during development. However, a recent study reported that pathogenic *FBRSL1* variants are associated with decreased expression of the chromatin regulators *BRPF1* and *KAT6A*, suggesting that *FBRSL1* plays a key role in regulating developmentally timed gene expression (Kastens, Berger-Santangelo et al. 2025). Our study, based on a single case, is not sufficiently powered to replicate these findings. Future functional work is needed to clarify this mechanism and may potentially lead to therapeutic targets.

Predicting the consequences of duplications is more challenging than deletions (Stankiewicz, Pursley et al. 2010). Deletions are likely to result in loss of function. Duplications may have multiple impacts: copy gain of triplosensitive genes; intragenic breakpoint causing protein truncation; intragenic breakpoint interfering with a splice site; or perturbation of distant regulatory elements. Determining which mechanisms are relevant is not straightforward. Our case emphasizes the importance of considering small duplications as pathogenic variants. Due to the small size (134 kb), the duplications in our patient would not be reported clinically on chromosomal microarray (CMA). Genome-wide short or long-read sequencing provides the opportunity to reveal these small copy number SVs.

GS is increasingly being used as a first-tier test in neonatal and pediatric intensive care units for specific indications where a genetic diagnosis could lead to an immediate change in management including for drug-resistant epilepsy (French, Delon et al. 2019; Seither, Thompson et al. 2024). Genome-wide analysis improves the diagnostic yield of genetic testing for neurologic and neurodevelopmental disorders (Palmer, Sachdev et al. 2021; Arnaud, Abi Warde et al. 2022; Johannesen, Tümer et al. 2023). Nevertheless, it has its limitations and challenges. Technological challenges include the limited ability to resolve complex SVs on short-read WGS, meaning long-read sequencing may be required, and knowledge gaps in functional interpretation of intronic and non-coding variants. Despite technological advances and new bioinformatic tools, we must also rely on comprehensive phenotyping to confidently match with genotype (Record and Reilly 2024). In our case the unique features of *FBRSL1-*related disorders including respiratory insufficiency, swallowing dysfunction, contractures, microcephaly, and heart malformations allowed us to confidently identify the disease-causing SV. Our findings emphasize the importance of incorporating robust SV analysis into GS workflows to identify small disease-causing SVs. Comparative phenotyping of patients is essential to understand the phenotypic spectrum of a disease-causing variant.

## Acknowledgments

We wish to acknowledge AUSiMED, and the Sima Weissman Foundation for their support by providing a scholarship to Dr Lital Cohen Vig for a clinical and research fellowship.

## Data availability statement

The raw data that support the findings of this study are not publicly available due to ethical restrictions and participant consent limitations. The dataset contains sensitive genomic information from an individual with a rare disease and their unaffected parents, for which informed consent was obtained only for use within the scope of this study. Requests for access to de-identified data will be considered on a case-by-case basis, subject to institutional ethics approval and a formal data sharing agreement. Enquiries should be directed to the corresponding author.

## Author Contributions

LCV and DK were responsible for patient recruitment and assessment. LCV and JEM drafted the first version of the manuscript and contributed equally as co–first authors. NS performed short variant analysis. JEM conducted structural variant analysis and pathogenic variant detection and interpretation. MFB performed repeat-expansion analysis. TW conducted ddPCR experiments. TW and JR performed the long-read DNA sequencing. MFB, LCV, and JEM carried out RNA-seq analysis. IES, MSH, MB, and SFB provided oversight, guidance, and editorial assistance.

